# Metagenomic sequencing as a clinical diagnostic tool for infectious diseases: a systematic review and meta-analysis

**DOI:** 10.1101/2020.03.30.20043901

**Authors:** Kumeren N. Govender, Teresa L. Street, Nicholas D. Sanderson, David W. Eyre

**Affiliations:** Nuffield Department of Medicine, John Radcliffe Hospital, University of Oxford, Oxford, UK; Big Data Institute, Nuffield Department of Population Health, University of Oxford, Oxford, UK

**Keywords:** infection, diagnosis, metagenomics, whole-genome sequencing

## Abstract

**Background:** Metagenomics has the potential to revolutionise infectious diseases diagnostics, from rapid species and antimicrobial resistance prediction, to finding unrecognised and sometimes untreated infections. Our aim was to summarise all literature on culture-independent metagenomic sequencing to describe the accuracy of species and antimicrobial resistance prediction and, describe the challenges and progress in the field.

**Methods:** We conducted a systematic review with meta-analysis from eligible studies retrieved from PubMed, Google Scholar and bioRxiv and, assessed risk of bias and quality using the QUADAS-2 tool. This study is registered with PROSPERO, number CRD42020163777.

**Findings:** We identified 36 studies, 22 of which used a species-agnostic approach to identify all possible pathogens. In these studies, the overall sensitivity and specificity of pathogen species detection were 88% (95%CI 81-92%) and 86% (95%CI 70-94%) respectively. Antimicrobial resistance prediction and comparison to phenotypic results was undertaken in six studies. Categorical agreement was 83% (95%CI 68-92%), very major (prediction sensitive, phenotype resistant) and major error (prediction resistant, phenotype sensitive) rates were 9% (95%CI 2-27%) and 1% (95%CI 0-20%) respectively. We report limited use of negative controls in studies 61% (22/36) which contribute to a major challenge of discriminating true pathogens from contamination, where there is no convergence on methodology. More efficient human DNA depletion methods are required as a median of 79% (IQR 62-96) [Range 7-98] of sequences were classified as human despite laboratory depletion techniques. The median time from sample to result was 23·5 hours (7-31) [4-144], with sequencing time accounting 10 hours (4·8-16) [1-16]. The average reported consumables cost per sample ranged from $128 to $685.

**Interpretation:** The science and regulatory environment are rapidly developing, and its role as a routine test or test of last resort still needs to be determined, however it is likely that clinical metagenomics will be an increasing part of the clinician’s armamentarium to diagnose infectious diseases in the near future.

**Funding:** None.

## Introduction

The term metagenomics first appeared in 1998 which referenced the idea that a collection of genes could be sequenced in a single sample without the need for isolation or lab cultivation of a specific species.^1^ Driven by advances in sequencing technology, the study of metagenomics primarily encompasses infectious disease diagnostics, microbiome analysis and oncological applications.^2^

In infectious diseases, metagenomics has the potential to revolutionise the field and provide rapid and precise diagnostics for patients, in place of traditional culture-based microbiology which has remained fundamentally very similar for much of the last century. The complete metagenomic pipeline from sample collection to report is shown in Figure 1. Traditionally most diagnostic microbiology depends on culture to identify the causative organism in an infection and to define antimicrobial susceptibilities. However, this process can take several days, or in the case of *Mycobacteria* and other slow growing organisms several weeks. While waiting for results patients are treated empirically, which may result in unnecessarily broad-spectrum treatment in some, and ineffective treatment in others. Additionally, culture is imperfectly sensitive, for example many patients with serious infection have negative blood cultures, and culture may be impaired by prior antimicrobial exposure.

**Figure 1.**
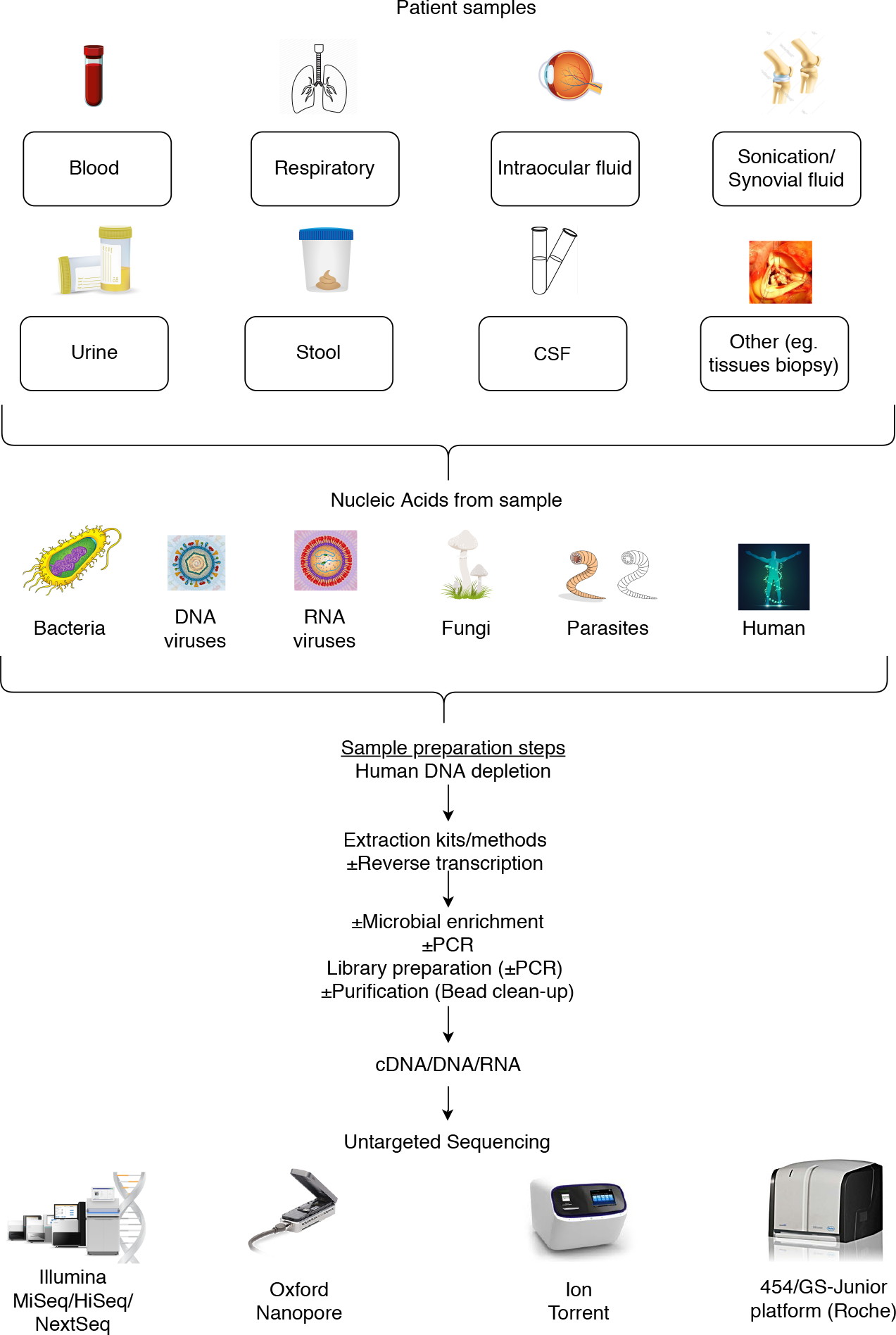
Metagenomic flow diagram. Patient samples undergo nucleic acid extraction. Laboratory methods may be used to reduce human DNA present, to improve pathogen sequencing efficacy. Reverse transcription may be utilised to convert RNA to cDNA for sequencing. Optional microbial enrichment is performed to further increase pathogen concentration. Random or specific PCR may be performed if a higher DNA concentration is required. Sequencer specific library preparation kits are used, which may include inherent PCR steps. Purification of the sample is often performed using bead-cleaning to remove inhibitors. Samples are then loaded onto the sequencer for sequencing.

Molecular diagnostics based on detection of antigens, antibodies or nucleic acid are increasingly used to complement culture-based diagnosis. For example, multiplex-PCR syndromic panels are able to identify common pathogens and antimicrobial resistance determinants within a few hours. However, these panels can only identify a restricted number of pathogens and resistance determinants and are therefore typically bespoke for a given clinical syndrome.

In contrast, emerging real-time sequencing technologies potentially offer a pathogen and clinical syndrome agnostic platform to identify any pathogens and all known resistance determinants present within hours. Studies have demonstrated that metagenomic pipelines can provide faster diagnoses than conventional culture-based methods which may hold crucial for critically unwell patients.^3^ This is particularly relevant for slow-growing organisms such as *Mycobacterium tuberculosis*; sequencing will likely play a key role in delivering faster diagnostics, quicker antimicrobial resistance prediction and enhanced outbreak investigation as part of the WHO’s End TB Strategy.^4^ Furthermore, metagenomics can yield diagnoses in challenging cases where conventional methods are uninformative, as in the case of a 29 year-old man incorrectly treated for tuberculous meningitis multiple times but found to have *Taenia solium* infection from metagenomic sequencing.^5^ Additionally the detection of pathogens from culture-negative samples may have implications in the clinical management in several other settings, e.g. in prosthetic joint infections, chronic wound infections and endocarditis.^6–8^

Metagenomic sequencing can also be used to identify previously described, as well as potentially novel, antimicrobial resistance determinants. Provided sufficient genetic context is present within a single sequence read, chromosomal resistance determinants can be matched to the species in which they occur. Determinants on mobile genetic elements, including plasmids, pose a greater challenge as these are harder to associate with a given host bacterial species, even with long-read sequencing technology. In contrast to culture-based techniques, obtaining results for a wide range of second- and third-line drugs requires minimal additional effort if sufficient sequence data for a given species is available together with knowledge of the genetic basis of resistance. Detection of mixed infections is also possible, where a subpopulation of resistant strains can be detected, that might be missed if only a limited number of colonies are sub-cultured for antimicrobial susceptibility testing.^9^

Whole-genome sequencing based on sequencing of isolates has substantively enhanced studies of infection transmission and outbreak investigations. Similar investigations can potentially be undertaken using metagenomic sequencing of clinical samples, e.g. during an outbreak of Shiga-toxigenic Escherichia coli (STEC) O104:H4 in Germany.^10^

Here, we review all current literature published on culture-independent metagenomic sequencing for infectious disease diagnostics and describe the specific challenges and progress in the field. We assess the accuracy of species identification and antimicrobial resistance prediction. We also describe the varying approaches taken to address specific challenges, including sample preparation involving human DNA depletion, unbiased DNA extraction methods often with limited pathogen DNA present, sequencing errors, bioinformatic approaches, databases and, contamination and quality control mechanisms.

## Methods

### Search strategy and selection criteria

We conducted a systematic literature review up to 27th February 2020. This systematic review was created in accordance with the Preferred Reporting System for Systematic Reviews and Meta-Analyses (PRISMA).^11^ We searched PubMed using a combination of phrases/keywords and using a set of relevant MeSH terms (Table 1). In addition, we performed a search with the same terms in Google Scholar and bioRxiv. Abstracts of studies identified from the literature search were screened using the inclusion and exclusion criteria specified in Table 2. Only studies based on metagenomic sequencing of all DNA present were included; studies based on 16S sequencing only were excluded. The reference lists of included manuscripts were also manually searched for relevant studies which may not have been identified during the literature search. Study inclusion was assessed by two independent reviewers and disagreements was judged by a third reviewer. This study has been registered with the PROSPERO prospective register of systematic reviews (reference CRD42020163777).

**Table 1.** Search strategy.

| Database | Query | Search terms | Number of items returned (27-02-20) |
| --- | --- | --- | --- |
| PubMed | 1 | "infection"[MeSH Terms] | 2556126 |
|  | 2 | "diagnosis"[MeSH Terms] | 8407178 |
|  | 3 | "whole genome sequencing"[MeSH Terms] OR "metagenomic"[MeSH Terms] | 44362 |
|  | 4 | "infection"[MeSH Terms]) AND "diagnosis"[MeSH Terms] AND ("whole genome sequencing"[MeSH Terms] OR "metagenomic"[MeSH Terms]) | 125 |

**Table 2.** Inclusion and exclusion criteria for this systematic review.

| Inclusion criteria | Exclusion criteria |
| --- | --- |
| Original manuscripts published in peer reviewed or pre-print journals | Conference abstracts. |
| Human studies | Animal and environmental studies |
| Clinical or spiked samples used | Case studies |
| Whole genome sequencing/metagenomics used | 16S rRNA gene sequence analysis |
|  | Microbiome studies |

Articles that utilised metagenomic sequencing as a tool for clinical diagnostics in infectious diseases were reviewed and relevant data was extracted where possible. The following data elements were extracted from all eligible articles: author, type of sample, range of pathogens identified, number of samples processed, nucleic acid extraction methodology, control samples used, sequencing technology used, classification software and database used, performance compared to standard methods, and cost and time of sample processing.

### Meta-analysis

Studies compared metagenomic sequencing to either standard microbiology, standard microbiology with additional tests or standard microbiology with clinical review. For the purpose of the meta-analysis, a pragmatic approach was adopted defining true positives based on each study’s best measure of the presence of infection with a specific species either by culture, molecular testing, clinical decision or a combination of these factors. Additional plausible species identified including polymicrobial infections were considered additional true pathogens and contributed to the numerator and denominator in the sensitivity analysis. Specificity was considered only in culture negative samples. Although it is plausible to consider that any sample can contain a false positive result by finding one or more species not identified by routine methods, studies differed considerably in reporting this.

We used the R meta package for the statistical analysis.^12^ We assessed methodologic quality and risk of bias using the QUADAS-2 tool for diagnostic accuracy studies.^13^ Outcomes, e.g. species identification sensitivity and specificity, are reported for each study with 95% confidence intervals (CIs), and pooled estimates provided using fixed or random-effects models, depending on the absence or presence of heterogeneity using the I2 test respectively. An I2 value >50% was considered heterogenous.^14^ Funnel plots and the Egger’s test were used to inspect for publication bias.^15^

## Results

The studies identified by the literature search are summarised in Figure 2: 132 manuscripts were screened, 96 were excluded for the reasons shown, leaving 36 eligible manuscripts for inclusion (Table 3).

**Table 3.** Characteristics of studies included in this systematic review.

| Study number | Study (Author and year) | Type of sample | Range of pathogens found | Positive samples | Negative samples | Nucleic acid extraction techniques | Sequencing techniques | Contamination | Cost per Sample |
| --- | --- | --- | --- | --- | --- | --- | --- | --- | --- |
| 1 | Wilson, M et al. (2017) | Cerebrospinal fluid | Fungi; parasites; viruses | 7 | Not found | Not found | Illumina next-generation sequencing | Yes, minimal | Not found |
| 2 | Miller, S et al. (2019) | Cerebrospinal fluid | Bacteria; fungi; parasites; DNA viruses; RNA viruses | 56 | 118 | EZ1 Virus Mini Kit v2.0 (Qiagen) | Illumina HiSeq | Yes | Not found |
| 3 | Xing, X et al. (2019) | Cerebrospinal fluid | Fungi | 16 | Not found | TIANamp Micro DNA Kit (Qiagen) | BGISEQ-500/50 platform | Not found | Not found |
| 4 | Wilson, M et al. (2019) | Cerebrospinal fluid | Bacteria; fungi; parasites; DNA viruses; RNA viruses | 58 | 149 | EZ1 Virus Mini Kit v2.0 (Qiagen) | Illumina HiSeq | Yes | Not found |
| 5 | Doan, T et al. (2016) | Intraocular fluid samples | Viruses; protozoa; fungi | 4 | 4 | Not found | Illumina HiSeq 2500 | Yes | Not found |
| 6 | Doan, T et al. (2017) | Intraocular fluid samples | Bacteria; fungi; parasites; DNA viruses | 31 | 36 | DNeasy Blood & Tissue Kit (Qiagen) | Illumina HiSeq 4000 | Not found | Not found |
| 7 | Anscombe, C et al. (2018) | Horse blood (Spiked with *E.coli* & *S. aureus*) | *E.coli*; *S. aureus* | n/a | n/a | Alkaline method | 454/GS-Junior platform (Roche) | Yes | Not found |
| 8 | Anson, L. et al. (2018) | Liquid blood culture (positive) | Gram-negative organisms; Staphylococcus sp. | 44 | Not found | BiOstic Bacteraemia; Amplex Hplex; MoLYsis Plus. | Illumina MiSeq; Oxford Nanopore Technologies (ONT) MinION | Yes | Not found |
| 9 | Gyarmati, P et al. (2016) | Whole blood | Bacteria; fungi; viruses | n/a | n/a | MoLYsis Complete5 kit (Molzym Life Science) | Illumina HiSeq 2500 | Yes | Not found |
| 10 | Hewitt, F et al. (2017) | Whole blood (spiked with fungi and bacteria) | Bacteria; fungi | n/a | n/a | AMPure XP 286 beads (BeckmanCoulter) | ONT MinION | Yes | Not found |
| 11 | Blauwkamp, T et al. (2019) | Plasma (cfDNA) | Bacteria; DNA viruses; fungi; eukaryotic parasites | 182 | 166 | Mag-Bind cfDNA Kit (Omega Biotek) | Illumina NextSeq 500 | Yes | Not found |
| 12 | Grumaz, S et al. (2016) | Plasma (cfDNA) | Gram-positive, gram-negative bacteria | n/a | n/a | Circulating Nucleic Acid Kit (Qiagen) | Illumina HiSeq 2500 | Yes | Not found |
| 13 | Hong, D et al. (2018) | Plasma (cfDNA) | Fungi | 9 | Not found | Magnetic bead-based method (Omega Biotek) | Illumina NextSeq 500 | Not found | Not found |
| 14 | McNaughton, A (2018) | Plasma | Hepatitis B virus | n/a | n/a | NucliSENS magnetic extraction system (bioMérieux) | ONT MinION; Illumina MiSeq | Not found | Not found |
| 15 | Parize, P et al. (2017) | Plasma | Bacteria; viruses | 19 | 82 | MoLYsis-5 reagents (Cat. No. D-321-050, Molzym). | Ion Proton (Life Technologies) | Yes | Not found |
| 16 | Greninger, A et al. (2015) | Plasma; whole blood; serum | Chikungunya virus, Ebola virus and hepatitis C virus | 4 | Not found | Direct-zol RNA MiniPrep Kit (Zymo Research); QIAamp Viral RNA kit (Qiagen); EZ1 Viral RNA kit (Qiagen) | Illumina MiSeq; ONT MinION | Yes | Not found |
| 17 | Horiba, K et al. (2018) | Plasma; serum | Bacteria; viruses | 12 | Not found | QIAamp UCP Pathogen Mini Kit (Qiagen) | Illumina HiSeq 2500 | Yes | Not found |
| 18 | Kafetzopoulou, L et al. (2018) | Plasma; serum | Chikungunya virus; Dengue viruses | n/a | n/a | QIAamp viral RNA kit (Qiagen) | Illumina MiSeq; ONT MinION | Not found | Not found |
| 19 | Sardi, S et al. (2016) | Serum | Chikungunya virus | 15 | Not found | Qiagen viral RNA minikit (Qiagen) | Illumina MiSeq | Yes, minimal | Not found |
| 20 | Del, C et al. (2018) | Serum | Hepatitis C virus | n/a | n/a | Total nucleic acid isolation (TNAI) kit (AmpliPrep System; Roche Diagnostics); QIAamp Viral RNA Mini Kit (Qiagen) | Illumina MiSeq; 454/GS-Junior platform (Roche) | Not found | Not found |
| 21 | Charalampous, T et al. (2018) | Respiratory samples | Bacteria | 33 | Not found | Saponin-based host DNA depletion | ONT MinION | Yes | \$130 per sample |
| 22 | Votintseva, A et al. (2016) | Respiratory samples | Tuberculosis | 39 | Not found | Ethanol precipitation method | Illumina MiSeq; MinION | Yes | £96-198 per sample |
| 23 | George, S et al. (2018) | Respiratory samples (Spiked with BCG) | BCG | n/a | n/a | AMPure XP beads (Beckman Coulter) | ONT MinION | Yes, minimal | Not found |
| 24 | Zoll, J et al. (2015) | Respiratory samples; Nasopharyngeal aspirates | Viral pathogens | 4 | 6 | DNeasy Blood kit (Qiagen) | Ion Torrent PGM system (Life Technologies) | No. | Not found |
| 25 | Farnaes, L et al (2019) | Respiratory samples (cfDNA) | Bacteria; fungi; parasites; DNA viruses; RNA viruses | 15 | Not found | Magnetic bead-based method (Omega Biotek) | Illumina NextSeq 500 | Not found | Not found |
| 26 | Street, T. et al. (2017) | Sonication/synovial fluid | Bacteria | 69 | Not found | Ethanol precipitation | Illumina MiSeq | Yes | Not found |
| 27 | Sanderson, N et al. (2018) | Sonication/synovial fluid | Bacteria | 7 | 2 | Ethanol precipitation | ONT MinION | Yes | Not found |
| 28 | Matthew, T et al. (2018) | Sonication/synovial fluid | Bacteria | 146 | 67 | BiOstic Bacteraemia (Qiagen) | Illumina HiSeq 2500 | Yes | Not found |
| 29 | Ivy, M et al. (2018) | Sonication/synovial fluid | Bacteria; fungi | 82 | 60 | MoBio Bacteremia DNA isolation kit (Qiagen) | Illumina HiSeq 2500 | Yes, 1 | Not found |
| 30 | Ruppé, E et al. (2017) | Sonication/synovial fluid | Bacteria; fungi; parasites; DNA viruses; RNA viruses | 63 | Not found | Ultra-Deep Microbiome Prep kit (Molzym) | Illumina HiSeq 2500 | Yes, 91 | Not found |
| 31 | Cheng, J et al. (2019) | Heart valves | Bacteria | 41 | 7 | TIANamp Micro DNA kit (Tiangen) | Illumina HiSeq | Yes, 2 | Not found |
| 32 | Zhou, Y et al. (2016) | Stool samples | *Clostridium difficile* | 22 | 5 | NucliSens easyMag automated system (Biomérieux) | Illumina HiSeq | Yes | Not found |
| 33 | Hasman, H et al. (2014) | Urine | Bacteria | 19 | Not found | Easy-DNA kit (Invitrogen) | Ion Torrent PGM system (Life Technologies) | Yes | Not found |
| 34 | Graham, R et al. (2017) | Urine | *Neisseria gonorrhoeae* | 13 | Not found | MasterPure DNA extraction kit (Epicentre) | Ion Torrent PGM system (Life Technologies) | Not found | Not found |
| 35 | Janes, V et al. (2017) | Urine | Bacteria | 9 | 32 | NucliSENS easyMag platform (Biomérieux) | Ion Torrent Proton platform | Yes | €116 per sample |
| 36 | Schmidt, K et al. (2017) | Urine (Some samples spiked with MDR strain) | *Escherichia coli* | 10 | Not found | MoLYsis Basic 5 Kit (MoLYsis) | ONT MinION; Illumina HiSeq 2000 | Not found | Not found |

**Figure 2.**
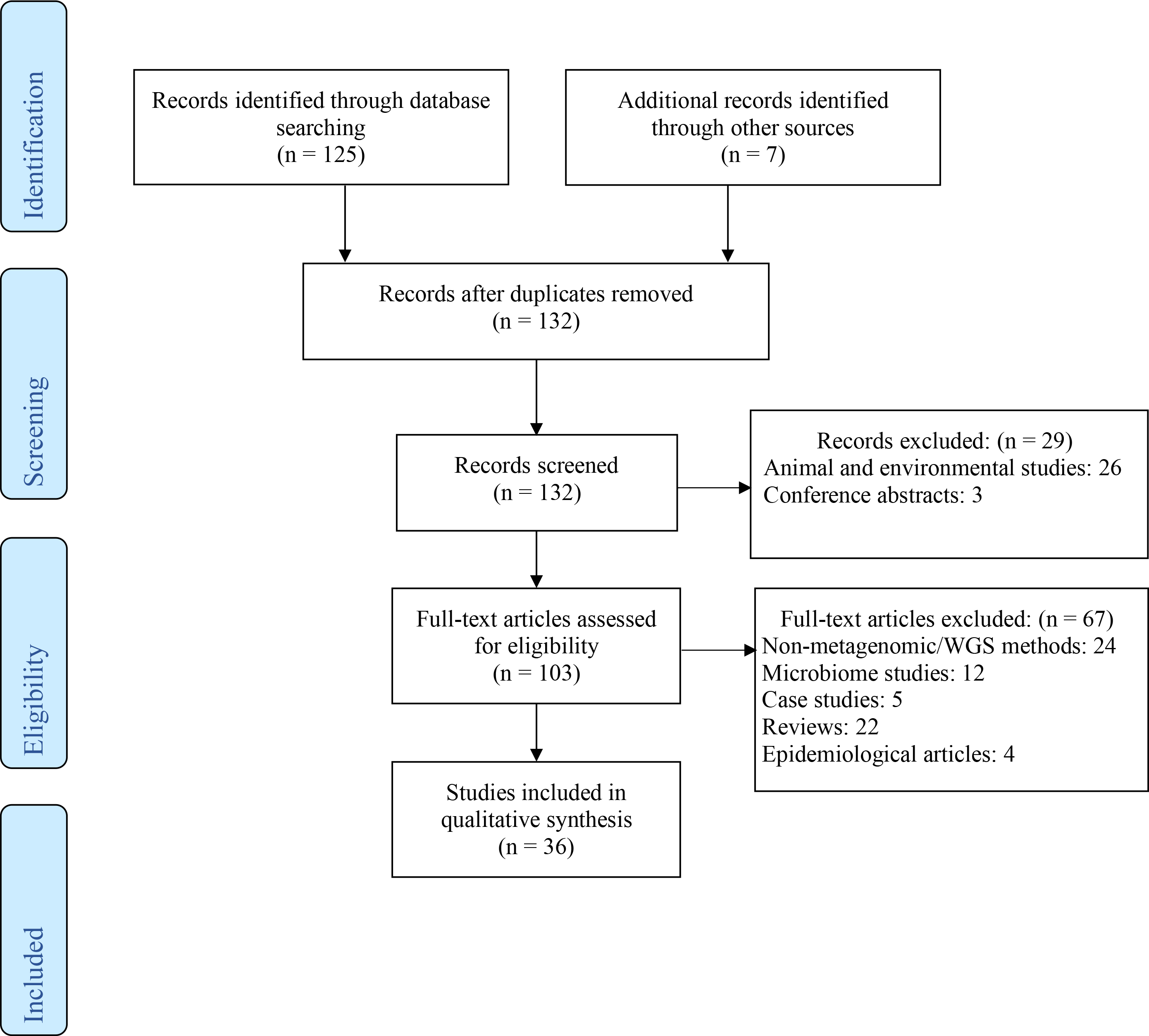
Flow chart showing study selection. 132 manuscripts were screened, 96 were excluded for the reasons shown, leaving 36 eligible manuscripts for inclusion.

### Clinical samples and range of pathogens

Of the 36 eligible studies identified, 14 (39%) undertook direct-from-sample sequencing of blood or positive blood cultures; three used cell-free DNA from plasma.^16–18^ Of these 14 studies, five focused on specific viruses, i.e. Hepatitis B and C, Chikungunya and Dengue virus;^19–23^ one focused specifically on fungal infections;^18^ and eight on bacterial infections.^3,16,17,24–28^ Five (14%) studies sequenced respiratory samples for bacteria,^29,30^ *M. tuberculosis*,^31^ spiked BCG samples^32^ and viral pathogens.^33^ Another four (11%) studies sequenced urine samples for bacteria^34,35^ and specifically for *Neisseria gonorrhoeae*^36^ and *Escherichia coli* including spiked multidrug resistant strains.^37^ Five (14%) studies analysed orthopaedic device sonication fluid and bone and joint samples for bacteria and fungi.^6,38–41^ Three (8%) studies analysed cerebrospinal fluid and two (6%) studies analysed intraocular fluid for multiple pathogens including bacteria, viruses, fungi and parasites,^5,42–44^ while one study (3%) analysed cerebrospinal fluid for fungi only.^45^ There were single studies of heart valve tissue for bacteria (3%) and stool samples specifically for *Clostridium difficile* (3%).^8,46^ The median number of samples sequenced per study was 30 (IQR 12-68) [Range 4-348].

### Methodology: nucleic acid extraction and sequencing library preparation

Several methods were used prior to DNA extraction to deplete human cells in 19/36 (53%) studies (Supplementary Table S2), including differential centrifugation^3,17,23,34^ or filtration with a 5μm membrane^28,38,39^ to remove human cells, based on their larger mass or size respectively, and the MolYsis Basic5 kit (Molzym, Bremen, Germany)^6,37,40^ which is reported to differentially lyse human cells.

Nucleic acid extraction kits and techniques were heterogeneous across studies (Supplementary Table S2). Most studies utilised commercial kits, primarily based on membrane/column methods, while non-commercial methods relied mostly on ethanol precipitation or lysis followed by bead-cleaning methods. Only one study compared the efficiency of different extraction kits.^3^ Seven (19%) studies performed a post-extraction enrichment step to facilitate enrichment of microbial DNA from clinical samples using the NEBNext® Microbiome DNA Enrichment Kit (New England Biolabs Inc., Ipswich, MA, USA) kit, which selectively binds and removes CpG-methylated host DNA.

14/36 (39%) studies used a DNA amplification method prior to library preparation to achieve required input DNA concentrations, which differed by the library preparation kit used. Of these studies, six studies used standard PCR amplification, five studies used multiple displacement amplification, two studies used Phi 29 rolling circle amplification and one study used primer extension preamplification PCR (Supplementary Table S2).

DNA clean-up with magnetic beads was used in 17/36 (47%) studies to purify extracted DNA prior to sequencing. For Illumina sequencing the Nextera XT DNA Library Preparation Kit was commonly used, with an input DNA requirement of 1 ng.^47^ Standard Oxford Nanopore methods were used in some studies, requiring 400 ng of DNA, or alternatively a combined PCR-amplification library preparation used in others, requiring 10 ng of DNA.^48^

### Methodology: controls

Studies were assessed for three types of controls, i) an internal or spiked control added to every sample processed to check for successful sequencing, ii) a negative control to allow contamination of reagents to be assessed and iii) a positive control that consisted of a known pathogen DNA. Three studies that tested spiked samples only were not assessed for internal controls.^24,26,32^ Of the remaining studies, 28/33 (85%) did not report the use of an internal control. Five studies reported internal control use including bacteriophages and synthetic DNA.^16,18,22,42,49^ Only 22/36 (61%) studies reported use of a negative control. The number of negative control samples varied from 0·7 to 2·9 (IQR 1·6-2·4) controls per every 10 samples processed. Only 6/36 (17%) studies reported use of a positive control, including using a mixture of 7 representative organisms and a single *Corynebacterium glutamicum* ATCC 13032 positive control.^6,16,18,30,42,49^

### Sequencing technologies

Illumina sequencing was used exclusively in 50% (18/36) studies (Supplementary Table S3). Four studies exclusively used Oxford Nanopore Technologies (ONT) MinION.^26,29,32,39^ Six studies compared the use of Illumina sequencing versus ONT MinION.^3,21–23,31,37^ Five studies sequenced on the ION Torrent/Proton.^27,33–36^ One study used the 454/GS-Junior platform (Roche),^24^ another the BGISEQ-500/50 platform (BGI-Tianjin, China)^45^ and a study evaluated the Versant HCV Genotype 2·0 assay (LiPA) versus the Illumina MiSeq and the 454/GS-Junior platform (Roche).^19^

### Bioinformatic approaches

Figure 3 outlines the bioinformatic workflows implemented. The two key components of each bioinformatic approach are the software used for classifying the species each read originated from and the associated sequence database. Bioinformatic pipelines were classified as either custom built (n=22), pre-existing (n=9) or commercial (n=5). The most commonly used classification software packages were Kraken (n=7), BLAST-related (n=7), SURPI (SNAP and RAPSearch) (n=3) and other/unspecified (n=19). The most commonly used databases originate from the NCBI including the NCBI RefSeq database (n=14), unspecified NCBI databases (n=7), NCBI Genbank (n=4) and NCBI reference genomes (n=2). Other studies developed bespoke databases (n=2), used proprietary ones (n=3) or used other specified databases (n=4) (Supplementary Table S3).

**Figure 3.**
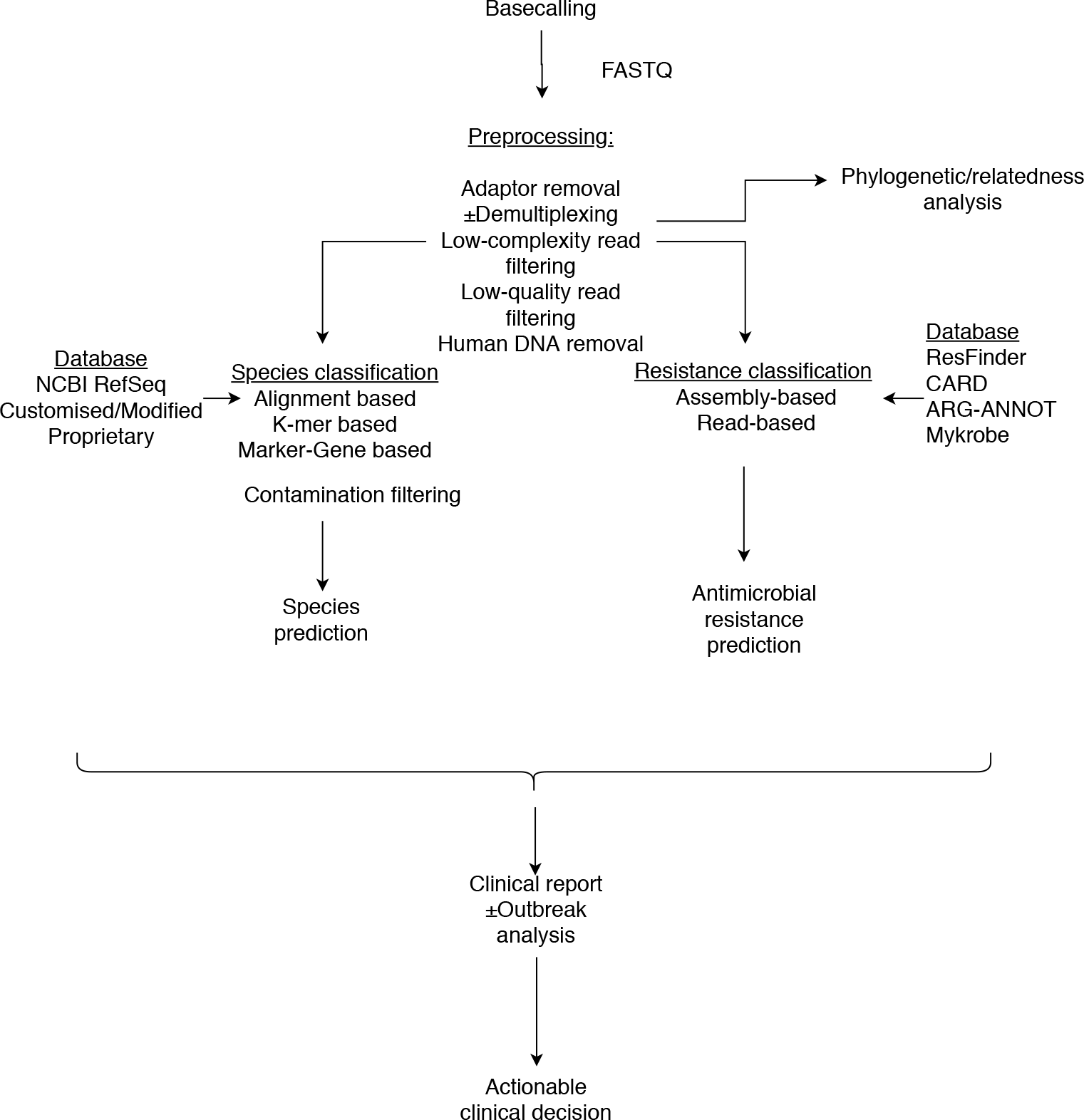
Metagenomic bioinformatic flow diagram. Raw data is basecalled to FASTQ files with corresponding quality scores. Preprocessing steps involve adaptor removal, demultiplexing, removing low-complexity and low-quality reads, and removing human reads. Species can then be classified using alignment, k-mer or marker-gene based software with appropriate databases. Contamination filtering may be performed thereafter. Resistance classification can be assembly- or read-based using the appropriate database. A relatedness/phylogenetic analysis may be performed for transmission analyses. A final clinical report ± outbreak analysis can be produced to provide potentially actionable clinical information.

### Assessing true species presence versus contamination

Various strategies were employed to identify true presence of an organism verses contamination (Figure 4). The most commonly used method include a parametric approach based on an assumed distribution for contaminating reads informed by numbers of reads in the control and other samples (n=6).^16–18,24,30,40^ Some studies (n=5) took an empirical approach to setting these absolute thresholds based on sequencing known negative and positive samples^3,6,39,42,49^ while, other studies developed composite measures to determine contamination using factors such as absolute and percentage reads, DNA quantity and relative importance of bacteria among others (n=5).^27–29,38,50^ Seven studies primarily identified contamination manually from known literature or if reported to be a contaminant previously in the local laboratory.^5,8,20,25,41,43,44^ In other studies (n=13) there was no description of how the presence of an organism was confirmed.^19,21–23,26,31–34,36,37,45,46^ Some studies (n=9) determined contamination without the use of a reported negative control by identifying known pathogens manually as described above (Figure 4).^3,20,22,26–29,34,50^ Therefore, by use of negative controls and other approaches, 26/36 (72%) studies report contamination of some degree while 9/36 (25%) studies had no details of contamination available. Only one study (3%) reported no contamination of two negative control samples out of 8 clinical samples in total.^33^

**Figure 4.**
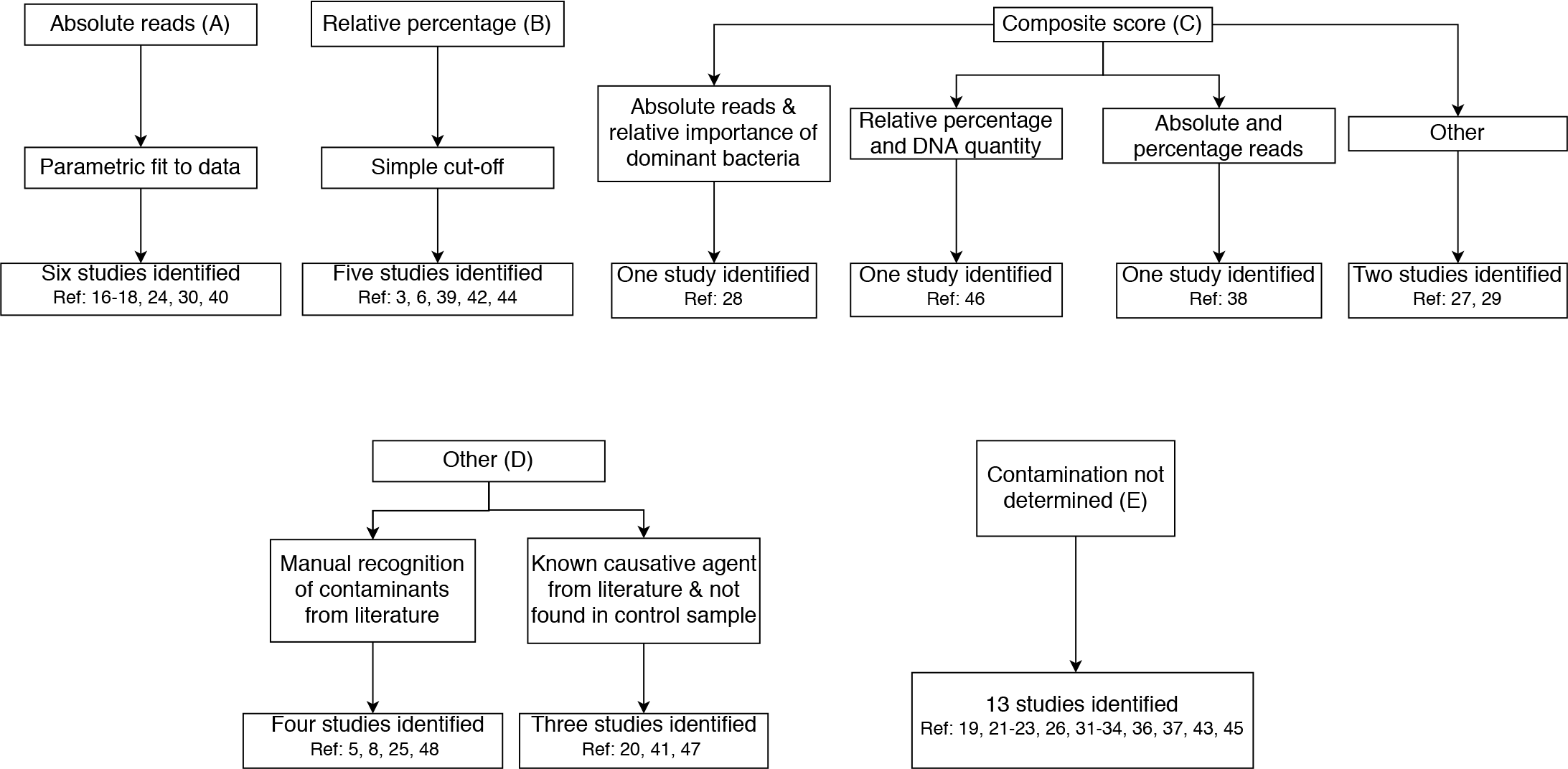
Common approaches to contamination control. Various strategies were utilised to identify true presence of an organism verses contamination. A: Parametric approach based on an assumed distribution for contaminating reads informed by numbers of reads in the control and other samples; B: Empirical approach to setting these absolute thresholds based on sequencing known negative and positive samples; C: Composite measures to determine contamination using factors such as absolute and percentage reads, DNA quantity and relative importance of bacteria among others; D: Manually from known literature or if reported to be a contaminant previously in the local laboratory; E: No description of how the presence of an organism was confirmed.

### Performance: species identification

The meta-analysis included 22/36 (61%) studies which contained a pathogen-agnostic workflow and allowed test sensitivity for detection of any organism to be analysed. Of these studies, 12/22 (55%) studies reported sufficient results from known negative samples to also assess specificity. We excluded 14 studies from the analysis of test sensitivity: 11 studies were not species-agnostic but instead focused on one or a few pathogens; and in three studies test sensitivity could not be determined (Supplementary Table S4).

We assessed the methodological quality of the 22 studies eligible for this meta-analysis using the QUADAS-2 tool (Figure 5).^13^ Considering the domains defined in the tool, patient selection, index test, reference standard, and flow and timing, 12/22 studies (54%) had low risk of bias in all; 7/22 (32%) had low risk and some concern of bias and 3/22 (14%) had high risk of bias in at least one domain (Supplementary Figure 2). In the applicability section, 13/22 studies (59%) had low concerns, 9/22 studies (41%) had some concern in one or more domains and no studies (0%) had high concern in any domain (Supplementary Table 5). Risk of bias was most commonly seen in the patient domain with 3/22 (14%) studies having high risk and 5/22 (23%) having some concern, all relating to selection of specific samples for study, rather than use of a random or consecutive sample. This was followed by the index test domain were 7/22 (32%) studies had some concern relating to the reporting of bioinformatic thresholds to assign a result as positive or negative.

**Figure 5.**
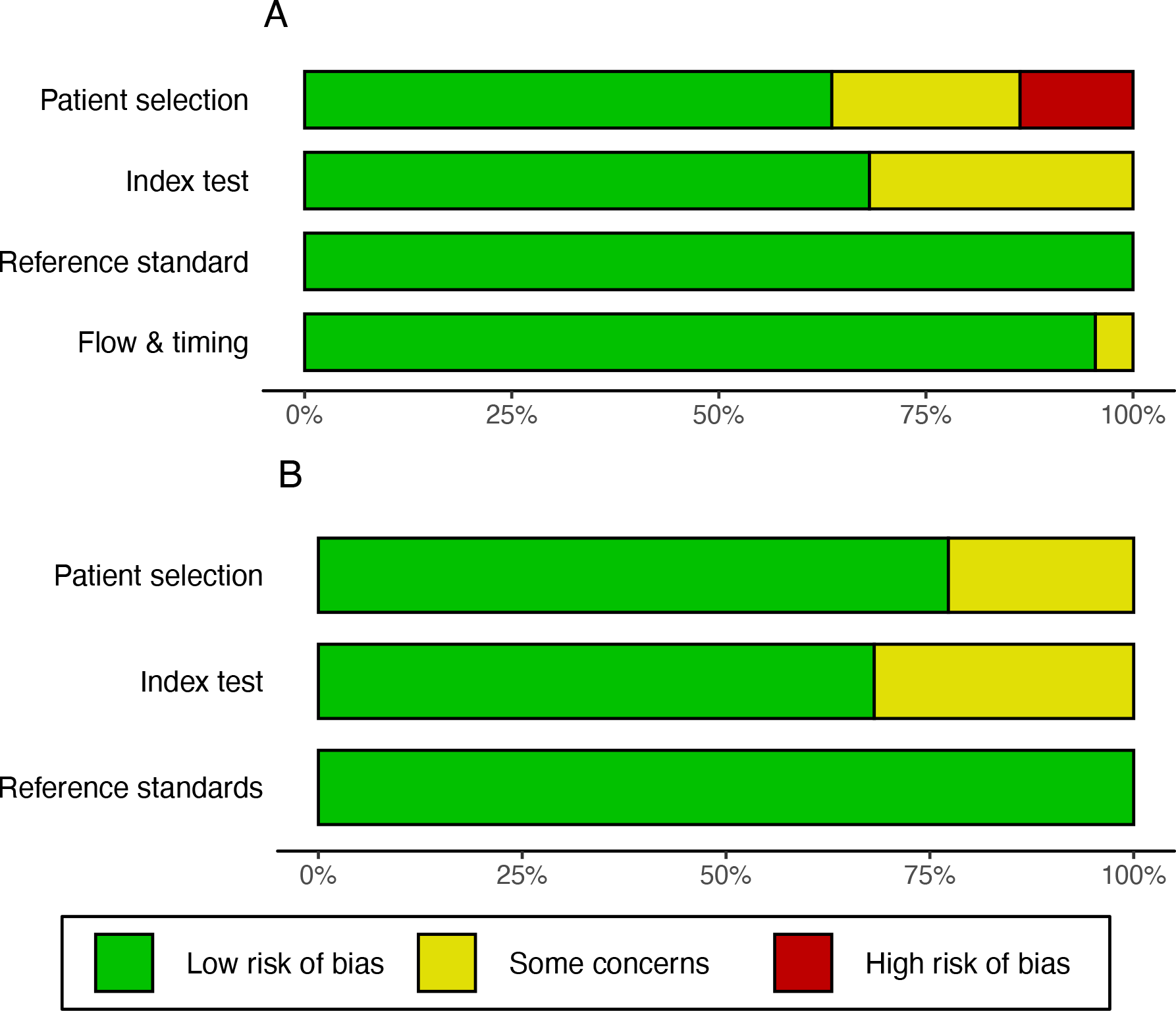
QUADAS-2 TOOL for the evaluation of primary research articles. The graphs present a summary of authors’ judgments with respect to each domain as percentages across all included articles. **A, Risk of bias of studies included in the meta-analysis**. This tool comprises 4 domains: patient selection, index test, reference standard, and flow and timing. Each domain is assessed in terms of risk of bias guided by signalling question listed in Supplementary Figure 1. **B, Applicability concerns of studies included in the meta-analysis**. This tool comprises 3 domains: patient selection, index test and reference standard. The findings for each included are listed in Supplementary Figure 2.

The pooled sensitivity of species identification across 22 studies was 88% (95%CI 81-92%) using a random effects model as there was heterogeneity between studies (I^2^ = 0·77, p<0·01). Sensitivity among sub-groups for each clinical sample type tested ranged from 82% to 98% (Figure 6). In the most widely investigated sample types CSF (n=121), blood (n=270) and orthopaedic (n=367) sensitivity was 0·84 (0·51-0·96), 0·85 (0·72-0·93) and 0·82 (0·72-0·88) respectively. Given fewer studies used negative controls or known negative samples, 12 studies were available to assess the pooled specificity, which was estimated to be 86% (95%CI 70-94%), again with marked heterogeneity (I^2^ = 0·94, p<0·01). Specificity among sub-groups ranged from 57% to 99% (Figure 7), and was 0·99 (0·96-0·99), 0·65 (0·59-0·71), 0·57 (0·32-0·79) in CSF, blood and orthopaedic samples respectively. 17 studies found a median of 6 (3-8) [1-62] additional pathogens with subsequent clinical interpretation varying throughout.

**Figure 6.**
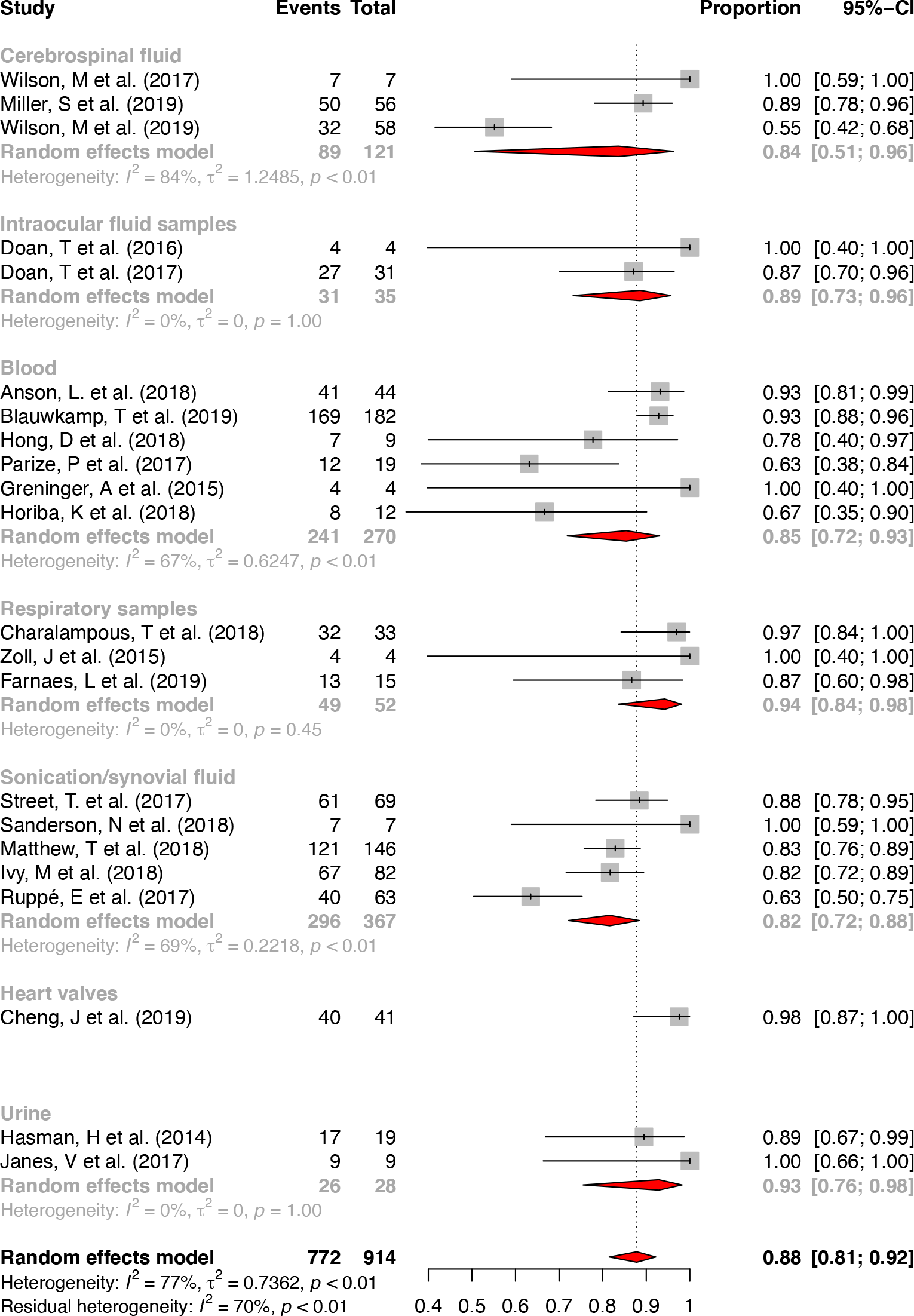
Pooled sensitivity of studies grouped by sample type. This analysis of sensitivity included 22/36 (61%) studies. 14 studies were excluded from the sensitivity analysis as 11 studies did not use species-agnostic samples and three studies where a sensitivity result could not be determined. Proportions were combined using a random intercept logistic regression model. A random effects model was used due to heterogeneity between studies.

**Figure 7.**
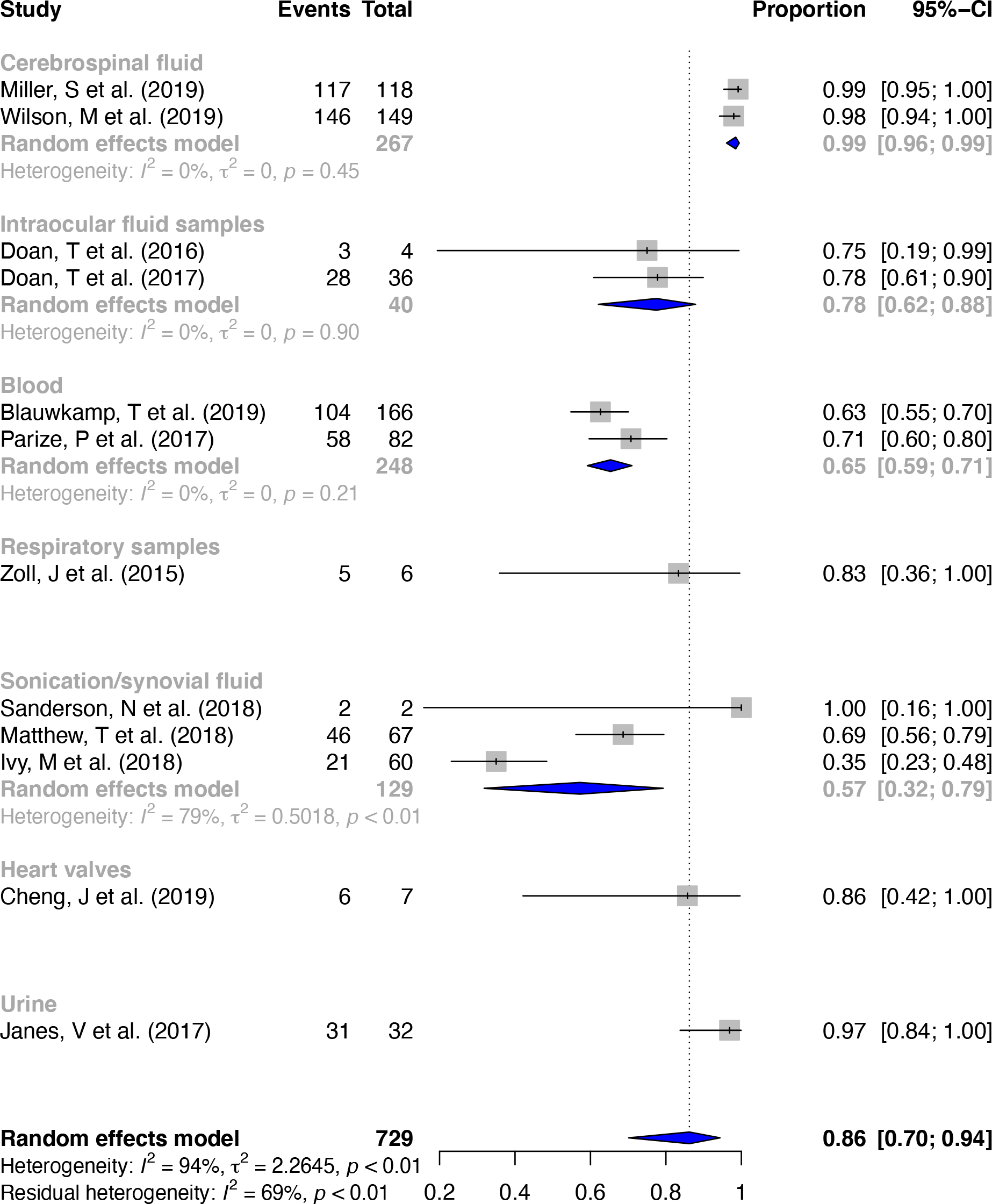
Pooled specificity of studies grouped by sample type. This specificity analysis included 12/22 (55%) studies that reported a specificity result. Proportions were combined using a random intercept logistic regression model. A random effects model was used due to heterogeneity between studies.

Funnel plots in both the sensitivity and specificity meta-analysis display evidence of heterogeneity present between studies. There is also evidence of asymmetry (Egger’s test: p=0·07 and p=0·04 respectively), but this favoured an excess of studies with lower performance (Figure 9.A/Figure 9.B).

**Figure 9.**
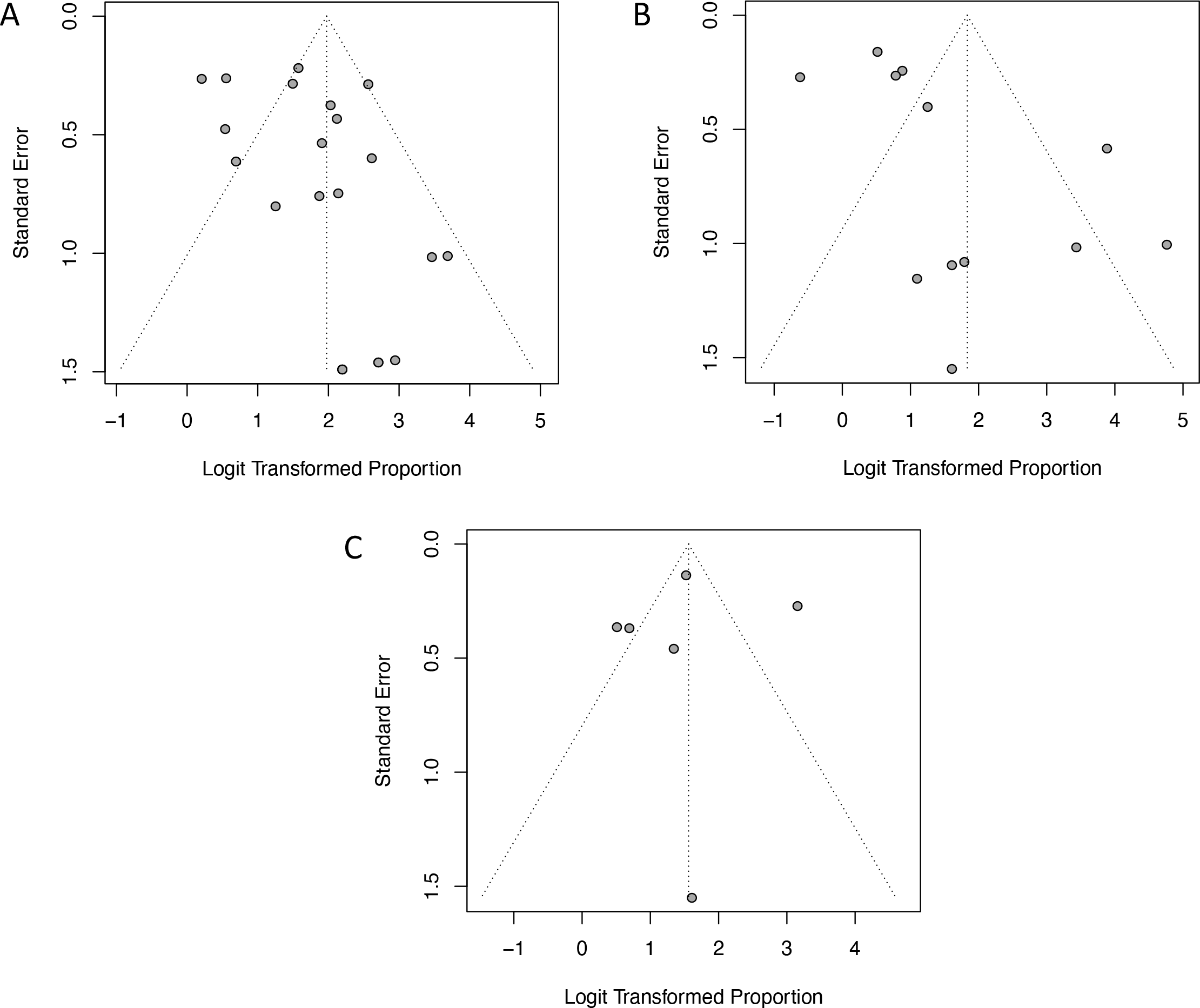
Funnel plots of meta-analyses. The vertical dotted line represents the summary estimate of the effect, while the other dotted lines represent the 95% confidence interval. **A**, Funnel plot of sensitivity meta-analysis. Egger’s test for asymmetry: p=0.07. **B**, Funnel plot of specificity meta-analysis. Egger’s test: p=0.04. **C**, Funnel plot of categorical agreement meta-analysis. Egger’s test not performed due to low sample size.

Using reported per-study means, a median 79% (IQR 62-96%) [Range 7-98%] of sequence data was classified as human DNA. Blood samples had a the highest median reported of 96% (79-96) [25-98], sonication and bone samples had a median of 89% (83-94) [76-98], respiratory samples had a median of 47% (47-47) [47-47] and urine samples had a median of 43% (25-60) [7-78].

### Performance: antimicrobial resistance prediction

15 studies attempted genotypic drug susceptibility prediction and compared results to either drug susceptibility phenotype (n=8), Illumina sequencing (n=1) or had no comparison (n=6). Of the 8 studies that compared results to drug susceptibility phenotypes, 6 studies were included in this meta-analysis that focused on species-agnostic samples (Supplementary Table S4). Performance depends on the prevalence of resistance and the specific pathogen-antimicrobial combinations tested. In these 6 eligible studies that compared phenotype and genotype, the median (IQR) [range] number of clinical samples per study was 21 (11-31) [1-39]. A median of 33 (30-266) [2-369] antibiotic predictions were performed per study. Performance was assessed as resistance prediction outcome over each combination of antibiotic and sample tested. All studies evaluated prediction of the correct resistance phenotype as a categorical value, i.e. sensitive or resistant. Categorical agreement rates were 83% (95% CI, 68% to 92%) across studies using a random effects model. The pooled estimate of very major error rates, defined as a phenotypic resistance not predicted by genotype was 9% (95% CI, 2% to 27%). The pooled major error rate, defined as predicted resistance by genotype but sensitive phenotype, was 1% (95% CI, 0% to 20%) (Figure 8). There was marked heterogeneity in all three metrics (I^2^ 0·89-0·94, all p<0·01; Figure 9.C).

**Figure 8.**
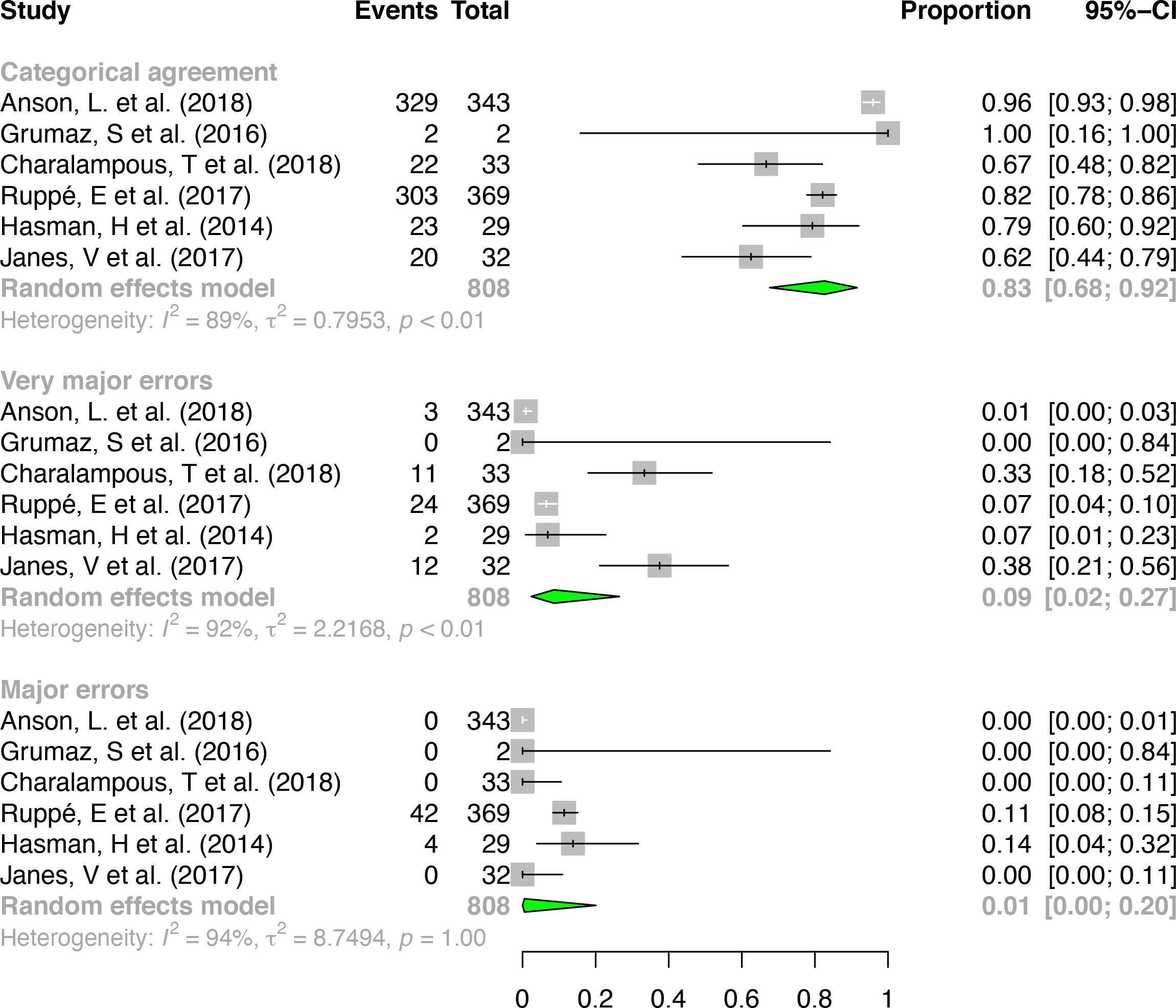
Pooled antimicrobial resistance prediction performance. This analysis included 6 studies that focused on species-agnostic samples and compared this to drug susceptibility phenotype. Proportions were combined using a random intercept logistic regression model. A random effects model was used due to heterogeneity between studies.

### Time of complete process

19/36 (53%) studies did not explicitly report the time taken from DNA extraction to speciation or antimicrobial susceptibility prediction. Figure 10 describes the time taken for sample processing in the remaining 17 studies. Despite studies using similar sequencing or DNA extraction methodologies there are differences in the times reported. Based on a single or mean time provided from each study, sample extraction/preparation took a median 3 hours (IQR 3-5) [range 0·5-5] and library preparation a median 3 hours (2-4) [0·1-15·5]. Reported sequencing took a median of 10 hours (4·8-16) [1-16], presumably because sequencing was terminated once a diagnostic result was reached in some cases, as some of these values are below the typical run times for many Illumina and Nanopore sequencing approaches. Where sequence analysis was undertaken in real-time using the Nanopore platform, first species prediction took 1 hour (1-4) [0·5-6] and AMR prediction took 3·5 hours (4·8-16) [2-5]. The median total time from sample extraction to speciation and/or AMR prediction was 23·5 hours (7-31) [4-144].

**Figure 10.**
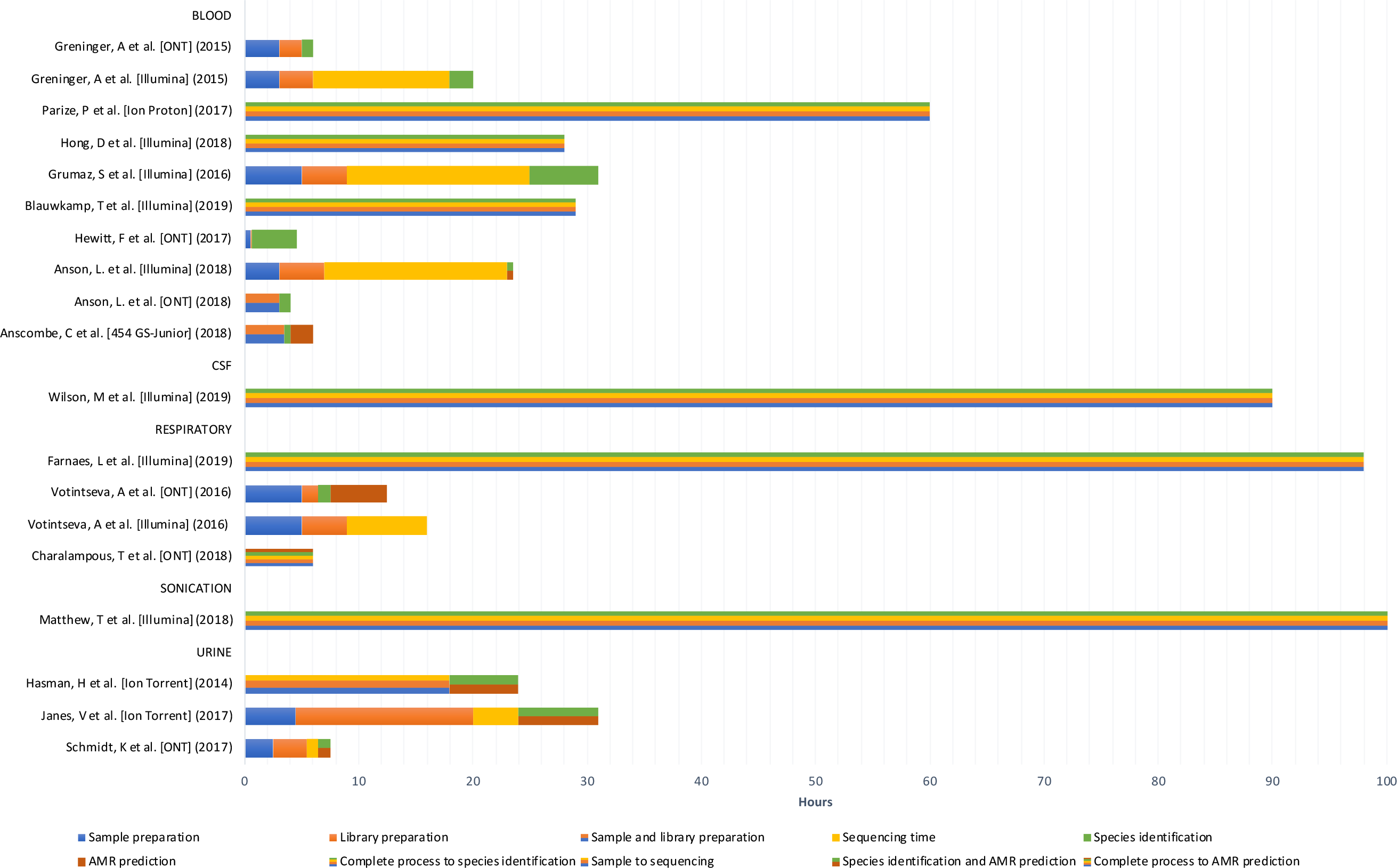
Time taken for sample processing grouped by sample type. The 17/36 (47%) studies shown below reported the time taken from DNA extraction to species identification or antimicrobial susceptibility prediction.

### Cost

Only three studies reported an average cost estimation per sample. Charalampous *et al*.^29^ estimated the reagent cost per sample to be $130 (Nanopore, 6 samples/run) or $546 (Nanopore, 1 sample/run). Votintseva *et al*.^31^ estimated the cost per sample to be $128 (MiSeq, 12 samples/run), $685 (R9 MinION, 1 sample/run) or $134-229 (R9·4 MinION, 3-5 samples/run). Janes *et al*.^35^ reported cost per sample to be $132 using the Ion Torrent Proton sequencer.

## Discussion

In this review we describe all studies to date assessing the performance of metagenomic sequencing as a tool for diagnosis of clinical infection. We identified 36 studies, 22 of which used metagenomic sequencing as an approach to identify all possible pathogens agnostically. In this group the overall sensitivity and specificity for pathogen species detection were 88% (95%CI 81-92%) and 86% (95%CI 70-94%) respectively.

These results reflect impressive progress in this field but fall short of the levels of accuracy demanded of many diagnostics used for routine patient care. However, it should be remembered that current gold standard culture-based microbiology is also imperfect, for example only 10-15% of blood cultures show growth with almost half growing contaminants.^51^ Potentially metagenomics may increase detection of pathogens, reflecting this, reported specificity may be reduced by additional true pathogens detected by metagenomics and not by culture. For example, a median of 6 (IQR 3-8) [range 1-62] additional pathogens were found per study (i.e. not classified as contaminants). Only two studies compared metagenomics to both standard microbiology testing and clinical review,^5,42^ however this approach may be needed in future studies to evaluate the plausibility of additional potential pathogens detected. Significant heterogeneity was seen between studies, reflecting the different study settings, sample types and metagenomic approaches taken.

While species identification is useful, rapid and precise antimicrobial resistance prediction, that is robustly linked to specific pathogens, will ultimately be required for mainstream adoption of clinical metagenomics. Although the categorical agreement rate with phenotyping was 83% (95%CI 68-92%), major and very major error rates were 1% (0-20%) and 9% (2-27%) respectively. Despite the major error rate being within the current regulatory threshold, the very major error rate was not. The FDA requires major errors <3% and the 95% CI on the very major error rate to be ≤7·5% at the upper limit and ≤1·5% at the lower limit.^52^ Several improvements are likely needed to improve performance, including increased yield of pathogen DNA, better understanding and cataloguing of the mechanisms underlying antimicrobial resistance, and finally optimised algorithms to detect these within metagenomic data. For example, large scale efforts have been made in TB to improve genomic prediction of antimicrobial resistance,^53^ these may need to be replicated for other organisms. Additionally, specific software for detection of resistance in metagenomic data, such as DeepARG, is under development.^54^

Quality control is imperative for reliable and valid metagenomic results and will be essential for regulatory approval for clinical applications. Negative controls are required to detect contamination and avoid false positive results. Positive and internal controls mitigate the risk of false negative results. Only 15% and 17% of studies used internal and positive controls. Similarly, only 61% used negative controls. We recommend all metagenomic studies consider carefully using all three forms of control.

The ability of metagenomics to detect microbes agnostically and at very low concentrations is a major advantage however this also comes with the ability to detect contaminants much more easily which poses a major challenge. Contamination may arise at all stages of the diagnostic workflow, including at sample collection, during transport and in the laboratory. As DNA is amplified prior to sequencing, contamination with previously amplified DNA in the laboratory is a major concern, as well as contamination of reagents and from other samples. Meticulous laboratory practice is required to minimise contamination risks, but this imposes a greater burden than is required for many culture-based techniques. However, despite this some residual contamination is likely; 26/36 (72%) studies reported contamination to some degree. Studies used various approaches to classify contamination which we have described in Figure 4, however there is no convergence on how to confirm the presence of infection bioinformatically and further research is required to interrogate the various methods on multiple datasets to determine the optimal approach.

The ideal nucleic acid extraction method in metagenomics would be representative, i.e. contain the same DNA composition as the original sample, and result in ample DNA yield. The choice of extraction method is dependent on the purpose of the study and generally involves a trade-off amongst efficacy, extraction time, ease of extraction, laboratory resources available, DNA yield, cost and/or human cell/DNA depletion requirements. There is a need to establish optimal extraction methods, only one study was found with a limited number of samples that compared different methods.^3^ Detailed information on library preparation methods can be found in a review by S. Head *et al*.^55^

For sequencing technology to be useful clinically it will require rapid turn-around times with high accuracy. It is yet unclear which sequencing platform will emerge as the ideal device for metagenomic sequencing of infectious diseases that has the potential to be scaled for everyday hospital use. For detailed information on sequencing platforms, a review is available by Goodwin S *et al*.^56^ Ion torrent and Illumina devices may be promising with high accuracy reads however the hurdle may lie with a 90-120 minute library preparation time and longer sequencing durations for both. In contrast, a quick 10-minute library preparation kit and real-time data generation provide an advantage to Oxford Nanopore technology which may be a game-changer for the diagnosis of infectious diseases in a clinical setting, but per read error rates are much higher with this platform at present.

As the quantity of microbial genetic data increases, bioinformatic approaches need to be optimised to handle large datasets of complex metagenomic data while minimising computing power and memory usage. A summary of the bioinformatic workflow for metagenomics is described in Figure 3. However, accredited workflows that can be reliably used by non-expert users will be needed for metagenomics to be adopted widely.

Human DNA present in metagenomic samples reduces the data generated on pathogens of interest, thereby increasing sequencing costs and decreasing sensitivity. A median 79% (IQR 62-96) [Range 7-98] of sequence reads were classified as human even after applying known human depletion laboratory techniques. Improved human DNA depletion is therefore required.

Numerous studies have shown that inadequate antimicrobial therapy is an independent risk factor for death among critically ill patients with severe infection.^57,58^ Furthermore, it has been demonstrated that a time interval of >1 day before initiation of appropriate antimicrobial therapy is associated with a 2-fold increase in the risk of mortality.^59^ Clinical metagenomics besides identifying fastidious organisms not identified by culture, may provide a species and antimicrobial result within 6-12 hours instead of the conventional 24-72 hour time-to-result which then can alter further antimicrobial therapy. Although the median time from sample to result in our review was 23·5 hours (IQR 7-31) [4-144], the bulk of time i.e. 10 hours (IQR 4·8-16) [Range 1-16] was spent on sequencing which may decline as real-time sequencing methods improve.

The average consumable cost per sample for clinical metagenomics is reported to range from $128 when samples are multiplexed up to $685 for processing a single sample at once. Although sequencing costs have dramatically reduced, the total reagent cost per sample continue to remain high, and these costs do not capture the skilled staff and specialised laboratory environment required for sequencing. In comparison, the average running cost for blood culture is estimated at <$50 per sample.^60^ However, as with blood cultures where <10% are positive, a key question is figuring out which patients are most likely to benefit from clinical metagenomics in clinical practice. A detailed cost-benefit analysis is needed to shed light on whether this technology will be beneficial and in what particular way.

Clinical laboratories are highly regulated with wide-ranging laboratory requirements that vary by jurisdiction, for example in the US FDA approval is required while in the EU a CE mark is required for diagnostic platforms. The FDA intends to regulate infectious disease next-generation sequencing devices as systems, including all the components necessary to generate a result. These components may include the sampling device, nucleic acid extraction, library preparation, reagents, sequencing, bioinformatic process and database to the final result. FDA has developed a database entitled “FDA-ARGOS” that supplies a set of validated regulatory-grade microbial genomic sequence entries as an alternative comparator method to existing culture or composite methods. Detailed information including quality control, validation and continuous monitoring may be found in the draft guidance by the FDA.^61^

Metagenomics may also offer enhanced capabilities during outbreak investigations.^62^ Studies have shown that phylogenetic tree construction using direct-from-sample metagenomics can yield highly similar results to sub-culture and whole genome sequencing of isolates^34,63^ Metagenomics may also potentially offer greater resolution if within host diversity can be accurately reconstructed and used for transmission inference.^64^ Furthermore, its agnostic ability allows detection of various unsuspected threats and even those from an accidental release of cultured or engineered organisms or from deliberate bioterror attacks.

Our study had several limitations. Firstly, the meta-analysis summary provided does not overcome problems that were inherent in the design and execution of the primary studies. Furthermore, it does not correct bias as a result of selective publication, where studies with better performance are more likely to be published. However, if anything, the asymmetry seen in reported results tended towards worse rather than better performance (Figure 9). Using the QUADAS-2 tool, the main risk of bias arose from selection of specific samples for study, rather than consecutive samples or a random subset which are recommended. Similarly, to avoid index test bias we recommend studies provide further clarity on bioinformatic procedures when determining a positive or negative result. Due to the heterogeneity between studies, we are not able to provide specific recommendations on the ideal metagenomics workflow but anticipate by bringing studies to date together this review will provide a mechanism for improving standards and designing better studies in future.

With further advances in extraction methods, sequencing technology and bioinformatic processes, the proficiency of clinical metagenomics is likely to improve while costs continue to decline. Culture-based diagnostics have remained the backbone of the microbiology lab for over a century;^65^ however clinical metagenomics has the potential to be the next frontier of clinical microbiology, not only enhancing our understanding of infectious diseases but also detecting polymicrobial infections and pathogens that are conventionally unculturable which may lead to more effective and narrow-spectrum treatment options reducing antimicrobial resistance and leaving the microbiome unchanged. It is highly likely that clinical metagenomics will be a part of the clinician’s armamentarium in the future to identify and guide therapy of infectious diseases.

#### Research in context

##### Evidence before this study

We searched PubMed, Google Scholar and bioRxiv using a combination of MeSH terms “infection”, “diagnosis” and “whole genome sequencing” OR “metagenomic” up to Feb 27, 2020, without language restriction and excluded case, microbiome and 16S rRNA studies (Table 2). No previous systematic reviews with meta-analysis were found. The widespread performance of clinical metagenomics is largely unknown, which is important for implementation of this technology at the bedside. We identified 36 studies, 22 of which used a species-agnostic approach to identify all possible pathogens. Quality and risk of bias was assessed using the QUADAS-2 tool.

##### Added value of this study

To address this uncertainty, we performed a meta-analysis of culture-independent metagenomic sequencing to describe the accuracy of species and antimicrobial resistance prediction and, describe the challenges and progress in the field. We found the overall sensitivity and specificity of pathogen species detection were 88% (95%CI 81-92%) and 86% (95%CI 70-94%) respectively. Concerning antimicrobial prediction, categorical agreement was 83% (95%CI 68-92%), very major (prediction sensitive, phenotype resistant) and major error (prediction resistant, phenotype sensitive) rates were 9% (95%CI 2-27%) and 1% (95%CI 0-20%) respectively. We also report the challenge of inadequate quality controls, contamination, regulation and cost among studies.

##### Implications of all the available evidence

While clinical metagenomics offers us a deeper and novel insight to treating infectious diseases, it may unlikely replace established diagnostics including PCR or multiplex assays in the next several years. We are likely to see some devices approved by the FDA in the near future which may first be used in reference laboratories particularly for complex undiagnosed infectious disease cases. In the long-term, clinical metagenomics may be established as a routine test for specific diseases and clinical scenarios such as encephalitis and meningitis to sepsis and pyrexia of unknown origin. The promise of an agnostic infectious disease diagnostic tool is revolutionary for clinical microbiology and has many advantages including detecting polymicrobial infections and pathogens that are conventionally unculturable which may lead to more effective and narrow-spectrum treatment options reducing antimicrobial resistance and leaving the microbiome unchanged. In the future clinical metagenomics is likely to be a part of the clinician’s armamentarium to diagnose and guide treatment of infectious diseases.

## Data Availability

Data available within the article or its supplementary materials.

## Contributors

KG and DWE were responsible for the study design, data acquisition, analysis, interpretation of results and drafting of the review. TS contributed to the study design and analysis. DWE, TS and NS critically revised the drafted review. All authors approved the final version of the review.

---

**Supplementary Figure 1.**
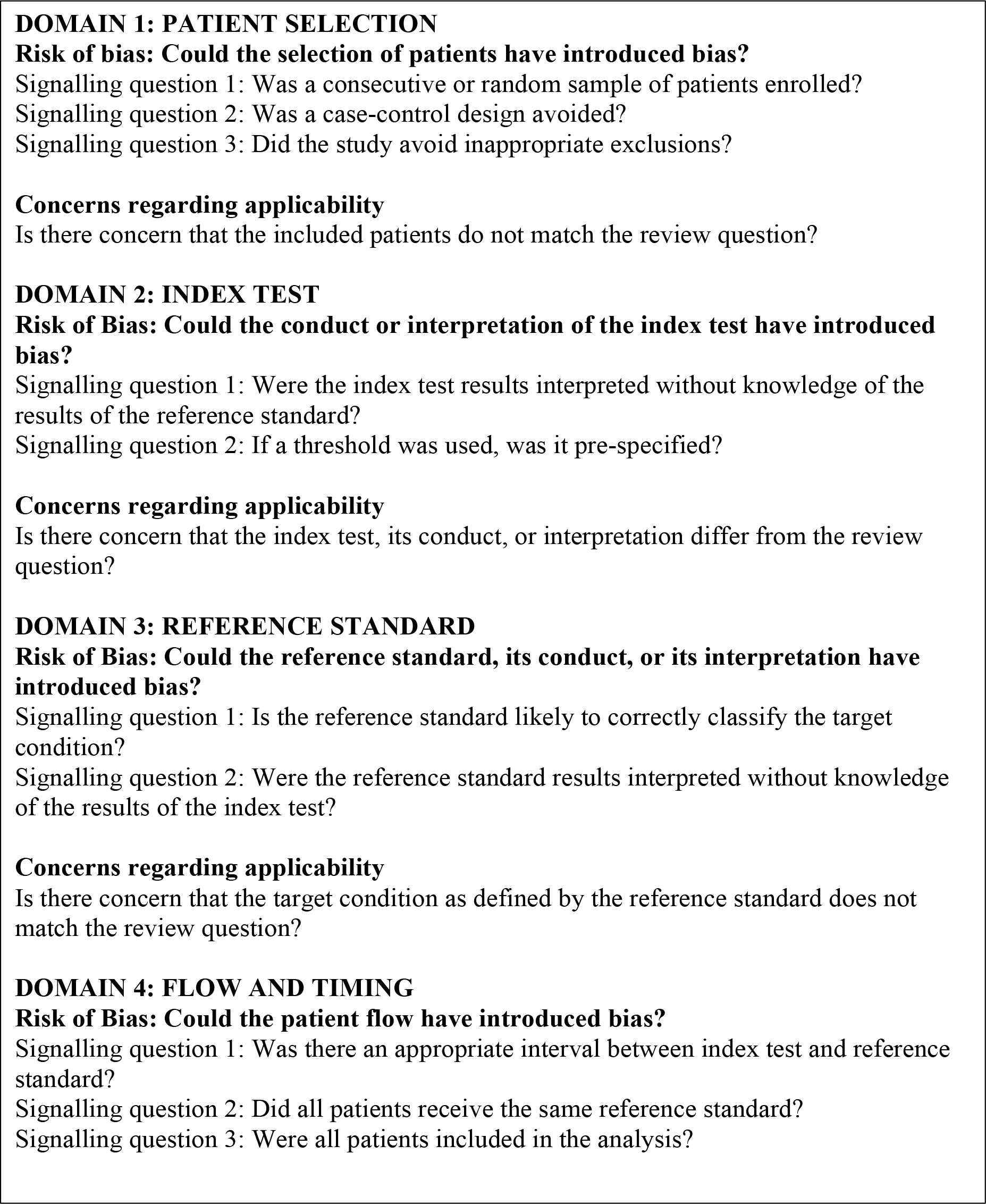
QUADAS-2 questions. Risk of bias is judged as “low”, “high”, or “unclear”. If all signalling questions for a domain are answered “yes” then risk of bias can be judged “low”. If any signalling question is answered “no” or “unclear” this flags the potential for bias. The “unclear” category was used only when insufficient data were reported to permit a judgment.

**Supplementary Figure 2.**
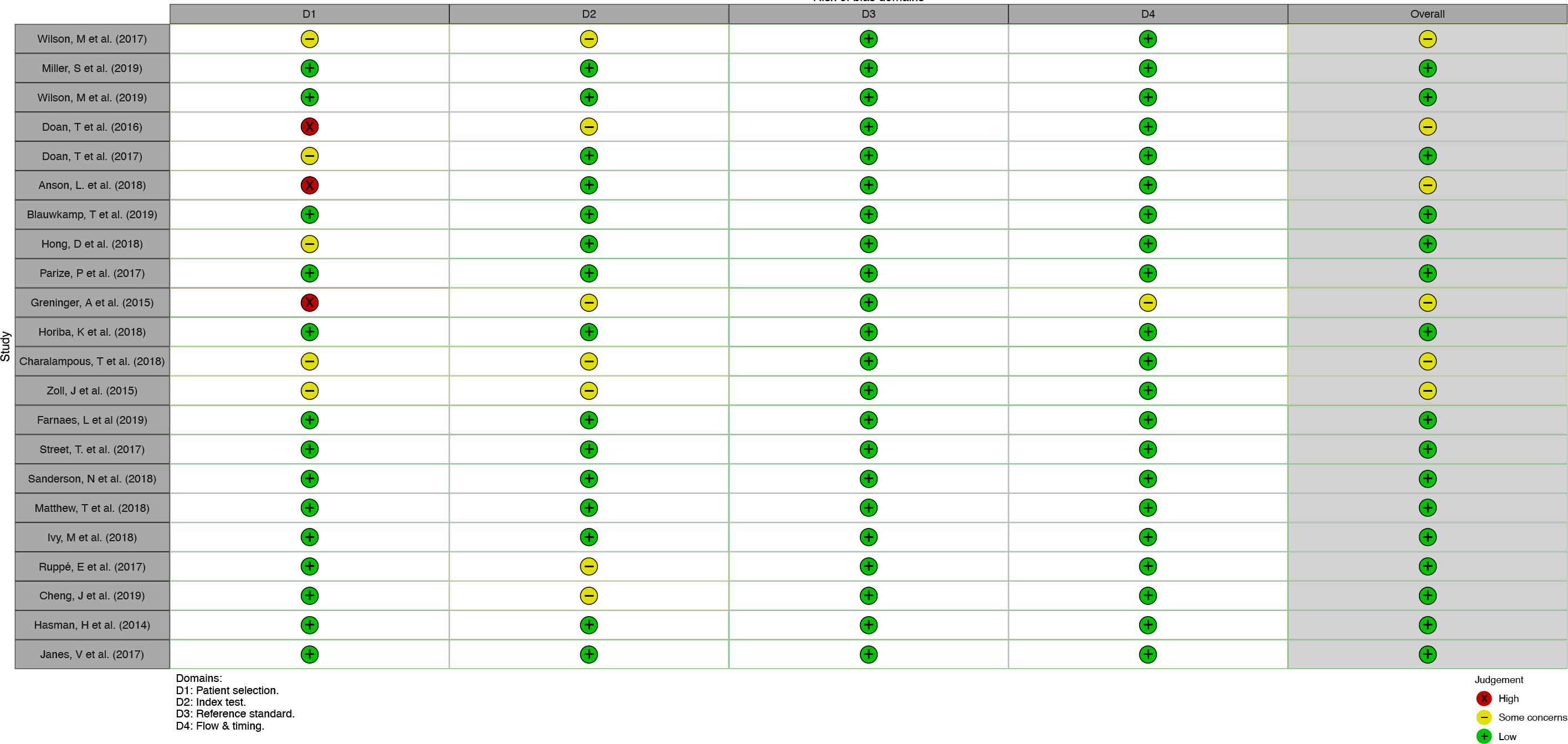
Traffic light plot of studies included in the meta-analysis. Individual study bias is shown below. For a detailed breakdown, refer to Supplementary Table 5 for signalling questions.

## Supplementary excel document

**Table S1. Laboratory methodology**

**Table S2. Bioinformatic details**

**Table S3. Performance of each study**

**Table S4. QUADAS-2 results**

